# Prevalence and spectrum of diabetic peripheral neuropathy and its correlation with insulin resistance - An experience from eastern India

**DOI:** 10.1101/2020.04.12.20056150

**Authors:** Manidipa Majumdar, Mainak Banerjee, Jinesh Sengupta, Soudeep Deb, Chanchal Kr. Jana, Biman Kanti Roy

**Author notes:** **Corresponding Author:** Manidipa Majumdar, Current Address: Sri Jayadeva Institute of Cardiovascular Sciences and Research, Bannerghatta main road, Bangalore, Karnataka, 560069. India.

## Abstract

**AIMS:** Diabetes mellitus is a public health problem worldwide, with diabetic neuropathy (DN) being a common complication. Studies indicate that, neurons can develop insulin resistance (IR) and cannot respond to the neurotrophic properties of insulin. Although studies exist on the relation between DN and glycemic exposure index (GE_i_), papers about correlation of DN with IR is rare. This study focused on the prevalence of neuropathies in DM patients and usefulness of IR as a marker of DN.

**METHODS:** A cross sectional observational study was done. All patients satisfying American Diabetes Association criteria and none of the exclusion criteria were included. Total sample size was 142. Main parameters studied were glycemic status, neurological examination and nerve conduction study findings. Dyck grading was used for severity of distal symmetric polyneuropathy (DSPN). For statistical analysis, logistic and ordinal logistic regressions were used as appropriate.

**RESULTS:** 34.5% of the sample had DN, the commonest type being DSPN (72.9%). The study population was equally divided in terms of gender and 88.7% were type 2 diabetic. About 62.5% neuropathic cases were asymptomatic. Occurrence of DN correlated significantly with duration of diabetes, FBS and IR. Age, when adjusted for other risk factors was not significantly correlated to neuropathy. The severity of DSPN correlated significantly with GE_i_ but not with IR.

**CONCLUSIONS:** The prevalence of neuropathy was found to be similar to earlier western studies. This paper further establishes IR as a significant predictor for existence of DN, but it may not affect the progress of the neuropathy.

## INTRODUCTION

### 1.1 BACKGROUND

Diabetes mellitus is a major public health problem both in developing and developed world. There is an alarming increase in the incidence and prevalence of diabetes mellitus particularly in Asian Indians [1], due to increased predilection for them to develop the disease. There is evidence showing that South Asians have greater insulin resistance even at comparable levels of total body fat percent and BMI, earlier impairments in β-cell function, greater propensity toward visceral fat deposition, even as neonates, and have lower levels of plasma adiponectin and higher levels of plasma leptin. More over Indians are undergoing a paradigm shift in social behavior where they are adopting western food habits and sedentary lifestyle, with growing urbanization. In 2000, India (31.7 million) had the highest number of people with diabetes mellitus in world followed by China (20.8 million) with the United States (17.7 million) in second and third place respectively [2], thus creating a large financial burden for control of diabetes and its related complications.

Diabetes is a complex metabolic disorder characterized by hyperglycemia and associated microvascular and macrovascular complications. Though macrovascular complications like cardiovascular diseases are major contributors to mortality, microvascular complications lead to prolonged morbidity, functional impairment and economic burden. Among microvascular complications, Diabetic neuropathy is a common and costly complication of both Type 1 and Type 2 Diabetes as well as the leading cause of non-traumatic lower limb amputations. Interestingly, it has been seen that Indians have a higher tendency to develop retinopathy and nephropathy compared to Caucasian population but a lower incidence of diabetic neuropathy and its associated complications [3][4]. In some studies, few of the factors attributed to such findings were shorter height, fewer pack-years smoked (among smokers), and higher transcutaneous oxygen levels (TCpO_2_). [5]

### 1.2 RISK FACTORS AND PATHOGENESIS

Peripheral neuropathy (PN) may be associated with varying combinations of weakness, autonomic changes and sensory changes. Diabetic Neuropathy (DN) is defined as ‘the presence of symptoms and/or signs of peripheral nerve dysfunction in people with diabetes after exclusion of other causes’ [6]. The spectrum ranges from a mild sensory disturbance as can be seen in most common form i.e. diabetic sensorimotor polyneuropathy (DSPN), to the debilitating pain and weakness of a diabetic lumbosacral radiculoplexus neuropathy. Many of these disorders of nerve appear to be separate conditions with different underlying mechanisms; some are directly caused by hyperglycemia whereas others are associated with diabetes [7].

HbA_1c_ values reflect overall glycaemic exposure over the prior 2-3 months [8]. The American Diabetes Association recommended treatment goal to prevent microvascular complications in type 2 diabetes mellitus patients is HbA_1c_ <7%, which is considered the standard for the monitoring of glycaemic control [9]. The onset of DN correlates with the duration of diabetes; 50% of patients develop DN after 25 years of diabetes[6]. Studies focussed on the glucose metabolic pathway suggest that overproduction of sorbitol and amino sugars due to activation of the polyol and hexosamine pathways, excess or inappropriate activation of protein kinase C (PKC) and accumulation of advanced glycation end-products contribute to the pathogenesis of DN.

Studies like the Diabetes Control and Complications Trial (DCCT) research group trial and UKPDS provide a clear connection between chronic hyperglycemia and the development of Diabetic Neuropathy [10] [11]. As an independent risk factor, plasma glucose level may be an important target among strategies to prevent or improve neuropathy. [12] [13] ‘Legacy effect’, the fact that glycemic memory of peripheral nerves over many years may determine the occurrence of neuropathies, has been discussed over the years [14]. The EURO-Diab group reported that blood glucose control, duration of diabetes, hyperlipidemia, hypertension, and smoking were all significant risk factors for the development of neuropathy in type 1 diabetic patients. [15]. The combined effect of duration of diabetes and glycaemic load has been studied in form of GE_i_ only a few studies which found positive correlation with occurrence of diabetic neuropathies, and predicted complications better than did individual components.

As a result of longterm hyperglycemia, a downstream metabolic cascade leads to peripheral nerve injury through enhanced advanced glycation end-products formation, an increased flux of the polyol pathway, aberrant release of cytokines, activation of protein kinase C and exaggerated oxidative stress, as well as other confounding factors. Although these metabolic changes are considered as the main theme for the development of diabetic microvascular complications, organ-specific biochemical and histological characteristics constitute discrete processes of neuropathy different from retinopathy or nephropathy. [16]

Insulin resistance is defined as a state of decreased responsiveness of target tissues to normal circulating levels of insulin and is the central feature of type 2 diabetes and Metabolic Syndrome[17] Several mechanisms have been described in the pathogenesis of DN mediated by insulin resistance. Studies in the last 10 years clearly suggest that insulin is a neurotrophic factor responsible for regulating neuronal growth, survival, differentiation and insulin receptors, signalling pathways are widely expressed in the nervous system. [18,19]

It was recently demonstrated that neurons indeed develop Insulin Resistance following hyperinsulinemia in a manner similar to that in metabolic tissues. [20] Chronic insulin stimulation was shown to induce insulin resistance in mouse Dorsal Root Ganglions, as evidenced by decreased activation of AKT and its downstream signaling pathway. This could attenuate the neurotrophic effects of insulin and subsequent development of neuropathy [21]. Insulin administration relieves painful DN [22] and reverses slowing of motor and sensory conduction velocities [23] in animal models of Type 1Diabetes, suggesting a direct role of insulin on neurons. Over the past decade, mitochondrial dysfunction has been shown to play a key role in the pathologic features of insulin resistance [24]. Excessive mitochondrial fission in cell bodies and neurons induced by insulin resistance [20] as well as hyperglycemia, may result in dysregulation of energy production, activation of caspase3 and subsequent dorsal root ganglion neuron injury [25,26].

Recent evidences indicate that similar to insulin-dependent metabolically active tissues such as fat and muscle, neurons also can develop Insulin Resistance and thus cannot respond to the neurotrophic properties of insulin, resulting in neuronal injury, subsequent dysfunction and diseases. Now if insulin resistance in diabetes is detected early and adequate steps are taken, it may be possible to significantly delay the occurrence of complications and there after their progression. A study demonstrated Insulin resistance is independently associated with peripheral and autonomic neuropathy in a group of Korean type 2 diabetic patients [27]. Although a lot of research works have already been carried out on the relation between diabetic neuropathies and HbA_1c_, duration of glycemia worldwide, the literature pertaining to the correlation of same with insulin resistance per se is not in abundance in India.

In this study we tried to find the prevalence and spectrum of presentation of diabetic peripheral neuropathies in Indian patients and whether their occurrence was dependent on traditional risk factors, glycemic exposure and insulin resistance.

## 2. METHODS

### 2.1 STUDY DESIGN AND SAMPLING

A cross-sectional observational study was conducted at R.G. Kar Medical College and Hospital in Kolkata, India. Data from cases were collected from both in-patient and diabetes clinic population, by simple random sampling, over a period of 6 months from January 2017 to June 2017.

Note that the optimal sample size required at 5% level of significance can be computed by *p*(1 − *p*)(*z*_0.025_)^2^/(0.05)^2^. Here, *z*_0.0.025_ denotes the 2.5% quantile of a standard normal distribution and its approximate value is 1.96. On the other hand, *p* is the true prevalence. Now, according to the National Urban Diabetes Survey (NUDS) [39], a population based study conducted in six metropolitan cities across India which recruited 11216 subjects aged 20 year and above representative of all socio-economic strata, the prevalence of diabetes mellitus in eastern India (Kolkata) is 11.2%. Thus, assuming 0.112 as the true prevalence, the optimal sample size is obtained as 153.

200 diabetic patients were selected by simple random sampling. Inclusion criteria for patients were as per the ADA criteria for diagnosing diabetes mellitus, and exclusion criteria included other known possible causes of peripheral neuropathies, patients with macrovascular complications, hepatic or renal dysfunction, previous history of neurologic disorders including stroke, smokers with regular smoking of one year or more, regular alcohol users for one year or more, malignancy, recent infection history, and autoimmune disease. The following flowchart (Figure 1) depicts the method of selection of subjects, using the above mentioned exclusion criteria, which finally yielded a sample size of 142 from whom we could gather all data of necessary variables.

**Figure 1:**
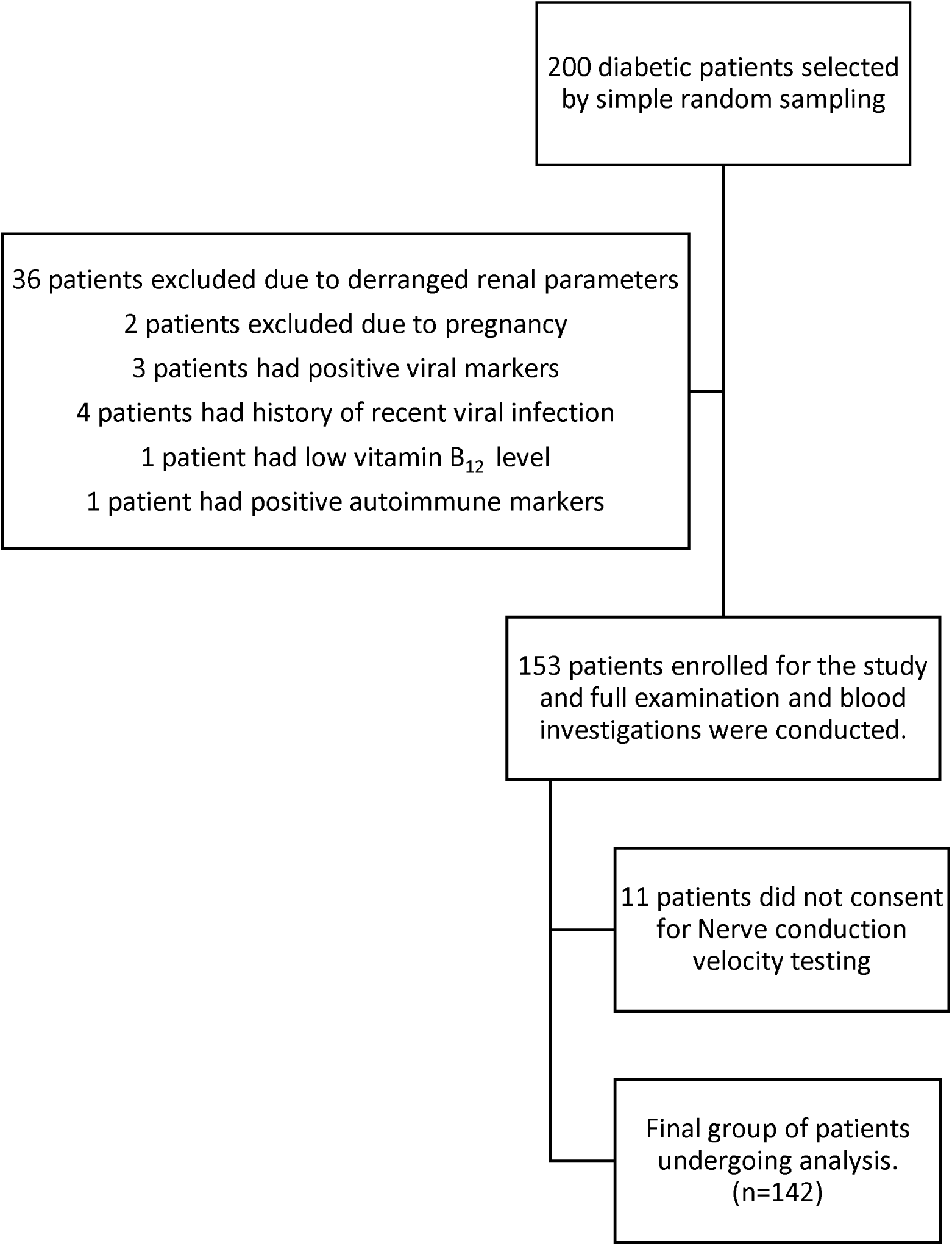
Method of selection of subjects in the study

### 2.2 METHOD OF DATA COLLECTION

Appropriate pre-tested medical history questionnaire was used to collect data on demographic characteristics like age, sex, as well as medical history suggestive of co-morbidities like hypertension, dyslipidaemia, drug use, etc. Apart from basic clinical examination, neurological examination was conducted where each patient was assessed for sensitivity, muscle strength; reflexes of the bilateral upper and lower extremities. Vibration sense was evaluated using a 128-Hz tuning fork and touch sensitivity with a 10 g monofilament. Nerve conduction studies were done for all patients. Main biochemical parameters assessed were fasting and post prandial plasma glucose, fasting c-peptide or insulin level, fasting lipid profile, renal function tests, liver function tests, routine hemogram, screening for viral markers, vitamin B_12_ and folic acid levels. ECG, and peripheral doppler were done to exclude ischaemic heart disease and peripheral vascular disease respectively.

### 2.3 OUTCOME DEFINITIONS

Diabetic Neuropathy was defined as ‘the presence of symptoms and/or signs of peripheral nerve dysfunction in people with diabetes after exclusion of other causes’[6]. Patients were broadly divided into 2 groups: (a) Diabetes mellitus patients with sensory and or motor neuropathy (b) Diabetes mellitus patients without sensory and or motor neuropathy. Subclinical neuropathy was defined as a person with no symptoms and signs but evidence of neuropathy on NCS. Asymptomatic neuropathy was defined as a person with no symptoms of neuropathy but with signs and or NCS abnormalities. The glycaemic exposure (GE) Index was calculated as follows: [28]

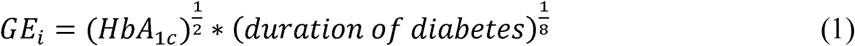

The homeostatic model assessment (HOMA) was the method used to quantify <u>insulin resistance.</u> First described under the name HOMA by Matthews *et al.* in 1985. Compared with the “gold” standard euglycemic clamp method for quantifying insulin resistance, quantification using modified HOMA-IR is convenient [29] and has been used in research in past.

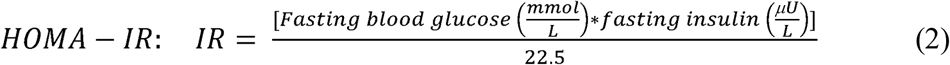

Now these parameters of both groups were compared to see if there’s a correlation between diabetic sensorimotor polyneuropathy and glycemic exposure or insulin resistance or both. Dyck grading was used to assess severity of polyneuropathy [30]: Grade 0: no abnormality of NC; Grade 1a: abnormality of NC, without symptoms or signs; Grade1b: NC abnormality of stage1a plus neurologic signs typical of DSPN but without neuropathy symptoms; Grade2a: NC abnormality of stage1a with or without signs (but if present, 2b) and with typical neuropathic symptoms; Grade 2b: NC abnormality of stage 1a, a moderate degree of weakness (i.e., 50%) of ankle dorsiflexion with or without neuropathy symptoms. Any correlation of these parameters with the severity of DSPN were also assessed if at all present. The Dyck grading was converted into an ordinal scale.

### 2.4 STATISTICAL ANALYSIS

In order to assess the effect of different regressors, we will use logistic regression model, since the response variable (presence/absence of neuropathy in a patient) is binary-type. Suppose, the response, hereafter denoted as *Y* has probability *p* of taking the value 1 (having neuropathy). Then, for regressors *X*_1_, *X*_2_,…, *X*_*p*_, the logistic model with a logit link is defined as

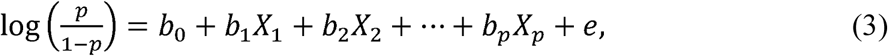

where *e* stands for a random error. The coefficients *b*_*1*_,…, *b*_*p*_ denote the effect of the different regressors in the model.

For any statistical analysis of this kind, appropriate variable selection method is also necessary. Multicollinearity, for example, is a phenomenon sometimes present in the data and can lead to erroneous results. It happens when two or more regressors are highly correlated. In light of that, as a first step of our statistical work, we will do a correlation analysis to remove any regressors showing signs of multicollinearity. Further, after the regression model is fit, we use stepwise variable selection method to identify most appropriate set of regressors for that relevant problem. This is done by choosing the sub model that displays least value of the Akaike Information Criterion (AIC).

On the other hand, we also analyze the effect of insulin resistance and glycemic exposure index on the degree of worsening of neuropathy, restricting ourselves to the cases with DSPN. Note that Dyck grading is an ordinal variable and hence, we will use ordinal logistic regression. The classes are denoted by 1a, 1b, 2a, 2b in order of increasing severity. Let us use 1 to 4 to denote these classes and *X* to denote the corresponding insulin resistance. Then, the ordinal logistic regression model for log-odds, for 1 = 1,2,3, is defined as

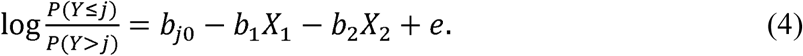

As before, *e* is a random error, *b*_*1*_ is the effect of insulin resistance, *b*_2_ is the effect of glycemic exposure index and the coefficients *b*_*j*0_ denote different intercepts for different classes.

## 3. RESULTS

### 3.1 DATA DESCRIPTION

The demographic characteristics of the study population are summarised in Table 1. First row indicates the observed prevalence of neuropathies. As described in table, half of the study populations were males, which describes no significant difference in gender distribution. 11.3% of the patients had type 1 diabetes which is expectedly significantly lower than the proportion of type 2 diabetes patients. 50.7% of the population had hypertension and 30.1% of the population had dyslipidaemia as co-morbidities. The prevalence is slightly less than that found in diabetic population in India but at par with diabetic population seen in larger international trials. The average HbA_1c_ of the study population was found to be 8.3 % which is higher than the usual target level of 7% (ADA 2016 guidelines), probably due to poor compliance. The mean fasting blood glucose was 168.19 mg/dl, and mean post prandial blood glucose was 239.50 mg/dl which shows that the mean values were considerably above target as per 2019 ADA guidelines. [31]

**Table 1:** Description of the study population

| VARIABLE | CATEGORY | NO. | MEAN | RANGE | STANDARD DEVIATION |
| --- | --- | --- | --- | --- | --- |
| Neuropathy | Present<br>Absent | 49 (34.5%)<br>93 (65.5%) | - | - | - |
| Sex | Male<br>Female | 71 (50.0%)<br>71 (50.0%) | - | - | - |
| Age (years) | - | - | 50.70 | (12, 82) | 16.53 |
| Type | T1D<br>T2D | 16 (11.3%)<br>126 (88.7%) | - | - | - |
| Duration of diabetes (years) | - | - | 9.02 | (1, 30) | 6.34 |
| Hypertension | With<br>Without | 72 (50.7%)<br>70 (49.3%) | - | - | - |
| Dyslipidemia | With<br>Without | 44 (31.0%)<br>98 (69.0%) | - | - | - |
| HbA<sub>1c</sub> (%) | - | - | 8.30 | (5.9, 12.6 ) | 1.43 |
| FBS (mg/dl) | - | - | 168.49 | (81, 347) | 58.04 |
| PPBS (mg/dl) | - | - | 239.51 | (110, 460) | 73.89 |
| Glycemic exposure index (GE<sub>i</sub>) | - | - | 3.66 | (2.43, 5.02) | 0.45 |
| Fasting C-peptide levels (ng/ml) | - | - | 3.3 | (0.7, 8.9) | 1.78 |
| Insulin resistance (IR) | - | - | 2.97 | (0.57, 7.69) | 1.69 |

### 3.2 PREVALENCE AND SPECTRUM OF NEUROPATHIES

The pattern of occurrence of neuropathies has been elucidated in Table 2. 33.8% of the patients included in the study population had evidence of neuropathy, which was greater than the prevalence found in some Indian studies [32] [33], but was at par with most large scale studies in western populations [34]. The commonest type of neuropathy seen was distal symmetrical sensorimotor polyneuropathy, which was seen in 72.9%, followed by entrapment neuropathies which were seen in 8.33% of cases.

**Table 2:** Prevalence of diabetic peripheral neuropathies. All percentages are computed based on total number of patients with neuropathy, which is 49.

| PREVALENCE | CATEGORY | SUB-CATEGORY | T1D | T2D | Total |
| --- | --- | --- | --- | --- | --- |
| Presentation | Clinical |  | 3 | 37 | 40 (81.6%) |
|  | Subclinical |  | 3 | 6 | 9 (18.4%) |
| Spectrum | DSPN |  | 5 | 30 | 35 (71.4%) |
|  | DSPN+EN |  | 0 | 3 | 3 (6.1%) |
|  | EN |  | 1 | 3 | 4 (8.2%) |
|  | SFN |  | 0 | 2 | 2 (4.1%) |
|  | MM |  | 0 | 2 | 2 (4.1%) |
|  | APN |  | 0 | 2 | 2 (4.1%) |
|  | CIDP |  | 0 | 1 | 1 (2.0%) |
| NERVE INVOLVEMENT | Upper Extremity | Median | 3 | 28 | 31 (63.3%) |
|  |  | Radial | 2 | 12 | 14 (28.6%) |
|  |  | Ulnar | 0 | 9 | 9 (18.4%) |
|  | Lower Extremity | Tibial | 3 | 28 | 31 (63.3%) |
|  |  | Peroneal | 2 | 27 | 29 (59.2%) |
|  |  | Sural | 4 | 31 | 35 (71.4%) |
|  | NCS inconclusive |  | 0 | 2 | 2 (4.1%) |

In addition, it is worth mention that 62.5% patients with neuropathy were asymptomatic. In type 1 diabetes patients, the proportion of subclinical neuropathies was higher (50%) than in type 2 diabetes patients (13.95%).

### 3.3 ASSOCIATION WITH RISK FACTORS

The study variables selected here are in accordance to the well-established role of glycaemic status on occurrence of microvascular complications as explained in major trials like DCCT. Insulin resistance, having a proven role in pathogenesis of diabetes as well as metabolic syndrome related disorders like pre diabetes, obesity, NAFLD, and diseases like PCOS, merits investigation in neuropathy cases also, as explained in section 1.2. Hypertension and dyslipidaemia were evaluated as ordinal variables as only their presence or absence was documented, not the severity. Both of these variables did not show any statistically significant association with neuropathy incidence, but inference cannot be drawn as absolute values of lipid levels and blood pressure would have been better metabolic parameters.

The data should be analysed separately for type 1 and type 2 diabetes because of significant differences in pathogenesis of the two diseases. However, since only 16 cases of type 1 diabetes were included in the study the sample size was inadequate for statistical tests. For completeness of this paper, we would like to mention some empirical analysis for that subset of the data. There are six type 1 diabetes patients with neuropathy, with equal occurrence across gender. The Pearson correlation coefficient of neuropathy with age, duration, fasting blood sugar (FBS), post prandial blood sugar (PPBS), HbA_1c_ and glycaemic index are found out to be 0.32, 0.76, 0.30, 0.04, −0.01 and 0.65, respectively. Now, the results from an extensive statistical analysis on type 2 diabetes patients are presented below.

We start with the set of regressors age, sex (male or female), duration, FBS, PPBS, glycaemic index, HbA_1c_ and insulin resistance. As mentioned before, we begin with a correlation analysis to identify multicollinearity and find no evidence of that. Next, a logistic model is applied with these regressors, where we include an interaction term for sex and duration as well. This was done to identify if the duration has different effects for males and females. Finally, a stepwise variable selection method is applied and subsequently, sex, interaction of sex and duration, FBS and insulin resistance are found out to be most appropriate set of regressors in the model. The results are presented in Table 3.

**Table 3:** Results from the logistic regression model of neuropathy on appropriate variables. * denotes significant effect on 5% level of significance.

| Regressors | Estimate | Standard error | p-value |
| --- | --- | --- | --- |
| (Intercept) | -7.92 | 1.52 | <0.001 * |
| Sex: Male | 1.13 | 1.09 | 0.300 |
| Duration for Male | 0.10 | 0.06 | 0.065 |
| Duration for Female | 0.19 | 0.07 | 0.003 * |
| FBS | 0.02 | 0.01 | 0.002 * |
| Insulin resistance | 0.71 | 0.71 | <0.001 * |

In order to avoid identifiability issues, coefficient corresponding to sex female is taken to be 0. Thus, the second row above indicates that males have a higher chance of having neuropathy, albeit it is not significant. The duration significantly increases the chance of having neuropathy for females, but the same is not true for males. FBS and insulin resistance, on the other hand, are both significant variables with a positive effect.

Lastly, we analyse the effect of glycaemic exposure index and insulin resistance on the Dyck grading. Restricting ourselves to the 33 patients with type 2 diabetes and DSPN, we use the ordinal logistic regression model. The results are provided in Table 4. Note that all intercept terms and the effect of glycaemic exposure index are significant. Insulin resistance, however, does not have a significant effect on the Dyck grading.

**Table 4:** Results from the ordinal logistic regression model of neuropathy on GE_i_ and IR for people with T2D and DSPN. * denotes significant effect on 5% level of significance.

| Variable | Estimate | Standard error | p-value |
| --- | --- | --- | --- |
| Intercept (1a:1b) | 9.68 | 3.81 | 0.01 * |
| Intercept (1b:2a) | 12.02 | 3.99 | <0.001 * |
| Intercept (2a:2b) | 13.36 | 4.12 | <0.001 * |
| Glycaemic exposure index | 2.65 | 0.90 | <0.001 * |
| Insulin resistance | 0.27 | 0.24 | 0.27 |

## 4. CONCLUSION

The prevalence of neuropathy, were found to be similar to western studies with DSPN being the most common presentation. Duration of diabetes, and fasting blood glucose positively correlated with occurrence of DN. It is however interesting that the variable selection method did not include age in the model while analysing the effect of different risk factors on the presence of diabetic neuropathy. In other words, the effect of age was found to be non-significant when other variables were considered. This finding is stark contrast to what was found in few other studies [35] [36]. It might be an indication that age has some co-dependence on duration of the disease and can determine insulin resistance, which proved to be a better predictor than either variable. In these previously mentioned studies insulin resistance was not investigated as a causative entity. Secondly, the effect of duration of diabetes was significantly associated with neuropathy akin to other studies which also showed significant effect of duration of diabetes [37] but in our study we found it significant only in females which might be due to genetic and metabolic variability between genders and should be investigated further. There was no significant difference in neuropathy occurrence between males and females unlike some studies which show male propensity to develop microvascular complications [38]. Insulin resistance was most significantly and independently related with occurrence of neuropathies, similar to the analysis of [27].

On the other hand, though there was increased occurrence of neuropathies in patients with increased IR, the severity of DSPN did not correlate with the extent of IR. This indicates that though higher insulin resistance increases the propensity to have diabetic neuropathy, its absolute value may not predict the severity of the disease, if we extrapolate our findings from the DSPN cases. Therefore, prospective follow-up studies are needed to evaluate its effect on progress of diabetic neuropathies. To the best of our knowledge, this aspect has not been studied earlier and is one of the interesting contributions of this paper.

Limitations of the study includes lack on investigation into autonomic neuropathies, and insulin resistance in type 1 diabetes patients. Since it is a hospital based study, selection bias might have been present while sampling. This study establishes IR as a risk factor for development of neuropathy. Studies can be conducted to extrapolate the hypothesis in other disease states with IR like pre-diabetes, non-diabetic obesity and poly cystic ovarian syndrome. Larger studies are needed in India to define geographical trends of prevalence and patterns of DN. Early focus on IR might be a crucial tool to prevent neuropathies.

## Data Availability

Data can be made available by authors on request.

## REFERENCES

1. Gujral UP, Pradeepa R, Weber MB, Narayan KV, Mohan V. Type 2 diabetes in South Asians: similarities and differences with white Caucasian and other populations. Annals of the New York Academy of Sciences. 2013 Apr;1281(1):51.

2. Kaveeshwar SA, Cornwall J. The current state of diabetes mellitus in India. The Australasian medical journal. 2014;7(1):45.

3. UK prospective diabetes study. XII: Differences between asian, afro-caribbean and white caucasian type 2 diabetic patients at diagnosis of diabetes. UK prospective diabetes study group. Diabet Med. 1994;11(7):670–677.

4. Kanaya AM, Adler N, Moffet HH, et al. Heterogeneity of diabetes outcomes among asians and pacific islanders in the US: The diabetes study of northern california (DISTANCE) Diabetes Care. 2011;34(4):930–937.

5. Abbott CA, Chaturvedi N, Malik RA, Salgami E, Yates AP, Pemberton PW, Boulton AJ. Explanations for the lower rates of diabetic neuropathy in Indian Asians versus Europeans. Diabetes Care. 2010 Jun 1;33(6):1325–30.

6. Boulton AJ, et al. Diabetic neuropathies: a statement by the American Diabetes Association. Diabetes Care. 2005;28:956–962.

7. Tracy JA, Dyck PJB. The Spectrum of Diabetic Neuropathies. Physical medicine and rehabilitation clinics of North America. 2008;19(1):1–v. doi:10.1016/j.pmr.2007.10.010.

8. Nathan DM, Buse JB, Davidson MB, Ferrannini E, Holman RR, Sherwin R, et al. Medical management of hyperglycemia in type 2 diabetes: a consensus algorithm for the initiation and adjustment of therapy: a consensus statement of the American Diabetes Association and the European Association for the Study of Diabetes. Diabetes Care 2009;32:193–203.

9. American Diabetes Association. Standards of medical care in diabetes--2012. Diabetes Care 2012;35 Suppl 1:S11–63.

10. Young MJ, Boulton AJ, MacLeod AF, Williams DR, Sonksen PH. A multicentre study of the prevalence of diabetic peripheral neuropathy in the United Kingdom hospital clinic population. Diabetologia. 1993 Feb 1;36(2):150–4.

11. DCCT Research Group. Factors in development of diabetic neuropathy: baseline analysis of neuropathy in feasibility phase of Diabetes Control and Complications Trial (DCCT). Diabetes. 1988 Apr 1;37(4):476–81.

12. Lu B, Hu J, Wen J, Zhang Z, Zhou L, Li Y, Hu R. Determination of peripheral neuropathy prevalence and associated factors in Chinese subjects with diabetes and pre-diabetes - ShangHai diabetic neuRopathy epidemiology and molecular genetics study (SH-DREAMS) PLoS One. 2013;8:e61053. doi: 10.1371/journal.pone.0061053.

13. Tesfaye S, Stevens LK, Stephenson JM, Fuller JH, Plater M, Ionescu-Tirgoviste C, Nuber A, Pozza G, Ward JD, EURODIAB IDDM Complications Study Group. Prevalence of diabetic peripheral neuropathy and its relation to glycaemic control and potential risk factors: the EURODIAB IDDM Complications Study. Diabetologia. 1996 Oct 1;39(11):1377–84.

14. Martin CL, Albers J, Herman WH, Cleary P, Waberski B, Greene DA, Stevens MJ, Feldman EL. Neuropathy among the diabetes control and complications trial cohort 8 years after trial completion. Diabetes care. 2006 Feb 1;29(2):340–4.

15. Tesfaye S, Chaturvedi N, Eaton SE, Ward JD, Manes C, Ionescu-Tirgoviste C, Witte DR, Fuller JH. Vascular risk factors and diabetic neuropathy. New England Journal of Medicine. 2005 Jan 27;352(4):341–50.

16. Yagihashi S, Mizukami H, Sugimoto K. Mechanism of diabetic neuropathy: where are we now and where to go?. Journal of diabetes investigation. 2011 Feb;2(1):18–32.

17. Sesti G. Pathophysiology of insulin resistance. Best Pract Res Clin Endocrinol Metab. 2006;20:665–79. doi: 10.1016/j.beem.2006.09.007.

18. Toth C, et al. Rescue and regeneration of injured peripheral nerve axons by intrathecal insulin. Neuroscience. 2006;139:429–449.

19. Xu QG et al. Insulin as an in vivo growth factor.Exp.Neurol. 2004;188:43–51.

20. Kim B, et al. Hyperinsulinemia induces insulin resistance in dorsal root ganglion neurons. Endocrinology. 2011;152:3638–3647.

21. Dunn TN, Adams SH. Relations between metabolic homeostasis, diet, and peripheral afferent neuron biology. Advances Nutr (Bethesda, Md) 2014;5:386–93. doi: 10.3945/an.113.005439.

22. Hoybergs YM, Meert TF. The effect of low-dose insulin on mechanical sensitivity and allodynia in type I diabetes neuropathy. Neurosci. Lett. 2007;417:149–154.

23. Brussee V, et al. Direct insulin signaling of neurons reverses diabetic neuropathy. Diabetes. 2004;53:1824–1830.

24. Turner N, Heilbronn LK. Is mitochondrial dysfunction a cause of insulin resistance? Trends Endocrinol Metab 2008;19:324–30.

25. Feldman EL. Oxidative stress and diabetic neuropathy: a new understanding of an old problem. J. Clin. Invest. 2003;111:431–433.

26. Vincent AM, et al. Mitochondrial biogenesis and fission in axons in cell culture and animal models of diabetic neuropathy. Acta Neuropathol. 2010;120:477–489.

27. Lee KO, Nam JS, Ahn CW, Hong JM, Kim SM, Sunwoo IN, Moon JS, Na SJ, Choi YC. Insulin resistance is independently associated with peripheral and autonomic neuropathy in Korean type 2 diabetic patients. Acta diabetologica. 2012 Apr 1;49(2):97–103.

28. Dyck PJ, Davies JL, Clark VM, Litchy WJ, Dyck PJ, Klein CJ, Rizza RA, Pach JM, Klein R, Larson TS, Melton LJ. Modeling chronic glycemic exposure variables as correlates and predictors of microvascular complications of diabetes. Diabetes Care. 2006 Oct 1;29(10):2282–8.

29. Henderson M, Rabasa-Lhoret R, Bastard JP, Chiasson JL, Baillargeon JP, Hanley JA, Lambert M. Measuring insulin sensitivity in youth: How do the different indices compare with the gold-standard method?. Diabetes & metabolism. 2011 Feb 1;37(1):72–8.

30. Dyck PJ. Detection, characterization, and staging of polyneuropathy: assessed in diabetics. Muscle & Nerve: Official Journal of the American Association of Electrodiagnostic Medicine. 1988 Jan;11(1):21–32.

31. American Diabetes Association. 6. Glycemic Targets: Standards of Medical Care in Diabetes-2019. Diabetes Care. 2019 Jan;42(Suppl 1):S61.

32. Pradeepa R, Rema M, Vignesh J, Deepa M, Deepa R, Mohan V. Prevalence and risk factors for diabetic neuropathy in an urban south Indian population: the Chennai Urban Rural Epidemiology Study (CURES-55). Diabetic Medicine. 2008 Apr;25(4):407–12.

33. Bansal D, Gudala K, Muthyala H, Esam HP, Nayakallu R, Bhansali A. Prevalence and risk factors of development of peripheral diabetic neuropathy in type 2 diabetes mellitus in a tertiary care setting. Journal of diabetes investigation. 2014 Nov;5(6):714–21.

34. Sun J, Wang Y, Zhang X, Zhu S, He H. Prevalence of peripheral neuropathy in patients with diabetes: A systematic review and meta-analysis. Primary Care Diabetes. 2020 Jan 6.

35. Booya F, Bandarian F, Larijani B, Pajouhi M, Nooraei M, Lotfi J. Potential risk factors for diabetic neuropathy: a case control study. BMC neurology. 2005 Dec 1;5(1):24.

36. Cheng WY, Jiang YD, Chuang LM, Huang CN, Heng LT, Wu HP, Tai TY, Lin BJ. Quantitative sensory testing and risk factors of diabetic sensory neuropathy. Journal of neurology. 1999 May 1;246(5):394–8.

37. Braffett BH, Gubitosi-Klug RA, Albers JW, Feldman EL, Martin CL, White NH, Orchard TJ, Lopes-Virella M, Lachin JM, Pop-Busui R, DCCT/EDIC Study Group. Risk Factors for Diabetic Peripheral Neuropathy and Cardiovascular Autonomic Neuropathy in the Diabetes Control and Complications Trial/Epidemiology of Diabetes Interventions and Complications (DCCT/EDIC) Study. Diabetes. 2020 Feb 12.

38. Singh SS, Roeters-van Lennep JE, Lemmers RF, van Herpt TT, Lieverse AG, Sijbrands EJ, van Hoek M. Sex difference in the incidence of microvascular complications in patients with type 2 diabetes mellitus: a prospective cohort study. Acta Diabetologica. 2020 Feb 5:1–8.

39. Ramachandran A, Snehalatha C, Kapur A, Vijay V, Mohan V, Das AK, Rao PV, Yajnik CS, Kumar KP, Nair JD, Diabetes Epidemiology Study Group in India (DESI. High prevalence of diabetes and impaired glucose tolerance in India: National Urban Diabetes Survey. Diabetologia. 2001 Sep 1;44(9):1094–101.

